# Rapidly Progressing Glaucoma: Clinical, Structural, and Socioeconomic Drivers of Treatment Escalation

**DOI:** 10.1101/2025.05.18.25327880

**Authors:** Lok Hin Lee, Yangyiran Xie, Chris Bradley, Jithin Yohannan

**Author notes:** **Corresponding Author and address for reprints:** Jithin Yohannan, MD, MPH Wilmer Eye Institute, 1800 Orleans Street, Baltimore, MD 21287. **Meeting Presentation:** This material has not been presented previously. **Financial Support:** Supported by grants from the National Institute of Health 1K23EY032204-03 and Research to Prevent Blindness Unrestricted Grant. **Proprietary Interest Statement:** Jithin Yohannan is a consultant for Abbvie, Topcon and Ivantis/Alcon. Chris Bradley is a consultant for Radius XR. The authors have no conflicting proprietary or financial interests related to this manuscript.

## Abstract

**Purpose:** To evaluate clinical and sociodemographic factors associated with selecting treatments in glaucoma patients with rapid visual field (VF) progression.

**Design:** Retrospective cohort study

**Participants:** 2,782 eyes from 1,812 adults with 5 or more 24-2 visual fields over five years and at least one optical coherence tomography (OCT) scan.

**Methods:** Rapid progressors were defined by mean deviation (MD) slopes worse than -1 dB/year. Demographic (age, gender, race), clinical (intraocular pressure (IOP), VF metrics, OCT measures), and socioeconomic (social vulnerability index, or SVI) variables were collected. Patients were categorized based on the most intensive treatment received in the first seven years: medical management, minimally invasive procedures (e.g., minimally invasive glaucoma surgery or laser), or aggressive procedures (e.g., filtering surgery or external ciliodestruction). Logistic regression was performed to identify demographic, clinical, and socioeconomic factors associated with treatment intensity.

**Main Outcome Measures:** Odds of treatment selection based on rapid VF progression

**Results:** Rapid progressors had significantly higher odds of receiving aggressive procedures (odds ratio [OR] 3.83, 95% confidence interval [CI] 2.56–5.74, p < 0.001) and any procedure (OR 3.15, 95% CI 2.28–4.35, p < 0.001), yet only 23% of rapid progressors underwent aggressive procedures in the first seven years. Among rapid progressors, worse MD and smaller rim area predicted aggressive procedures and higher IOP predicted any procedure. Higher SVI was associated with a reduced likelihood of receiving minimally invasive procedures among rapid progressors (OR 0.05, 95% CI 0.00–0.76, p = 0.031).

**Conclusion:** Although rapid progression was a strong predictor of aggressive procedures, fewer than one in four underwent aggressive IOP-lowering interventions. Baseline IOP and structural severity appeared to supersede VF progression in clinical decisions. Patients in areas of higher socioeconomic vulnerability were also less likely to receive less invasive procedures. Better integrating rates of functional decline and addressing socioeconomic barriers may help optimize care for rapidly progressing glaucoma patients.

## Introduction

Glaucoma remains one of the foremost causes of irreversible blindness worldwide and is anticipated to affect an expanding proportion of the aging population^1, 2^. Because the disease often progresses silently in its early stages, prompt detection and intervention are critical for preserving visual function and preventing long-term disability. Visual field (VF) testing is commonly employed to monitor disease progression, with the rate of change in mean deviation (MD) serving as a key measure of decline in visual function. Multiple studies suggest that approximately 5-10% of patients with glaucoma exhibit rapid progression, most commonly defined as eyes experiencing MD worsening exceeding -1 dB/year^3, 4, 5^.

A variety of clinical factors have been linked to faster rates of progression. Older age, elevated intraocular pressure (IOP), worse baseline MD, disc hemorrhage, and pseudoexfoliation have all been implicated in more rapid progression^6, 7^.

Beyond traditional ocular and systemic risk factors, social determinants of health have emerged as critical considerations in glaucoma care. Composite indices such as the Social Vulnerability Index (SVI) and Area Deprivation Index (ADI) incorporate multiple metrics—educational attainment, income, and housing stability, among others — have been used in previous studies on glaucoma to gauge community-level disadvantage^8, 9, 10^. Although higher levels of SVI or ADI may correlate with delayed access to treatment, more advanced disease at presentation and faster rates of glaucoma worsening^11^, some studies also suggest that individuals residing in high-SVI (more disadvantaged) areas might eventually obtain more aggressive interventions, reflecting complex and sometimes counterintuitive relationships between socioeconomic status and healthcare utilization^12^.

Within this complex landscape, treatment approaches can vary widely. Although aggressive procedures, such as tradition filtering surgery, has been shown to slow VF loss in glaucomatous patients^13^, there remain no clear criteria for escalating beyond medical, laser, or minimally invasive surgical interventions, and clinical practices differ widely. Understanding the drivers behind treatment decisions for aggressive surgery versus less invasive therapies, especially for rapidly progressing patients, can, therefore, assist in developing best practices and identifying ways to improve care.

Here, we aim to identify glaucoma patients who demonstrate rapid progression and examine the ocular, clinical, and socioeconomic factors that shape treatment strategies. Specifically, we explore whether these rapidly progressing individuals are likely to undergo aggressive procedures, such as filtering surgery, or whether they remain on medical treatment or receive less invasive procedures. We aim to clarify the underlying contributors to treatment escalation and to highlight potential gaps in care for patients who, despite substantial functional decline, may not receive robust intervention.

## Methods

### Study Population

This study was conducted in accordance with the tenets of the Declaration of Helsinki and was approved by the Johns Hopkins University School of Medicine Institutional Review Board. As this was a retrospective study using data collected in routine clinical practice, the need for informed consent was waived by the Institutional Review Board.

We included all adult patient eyes with glaucoma or glaucoma suspect diagnosis based on ICD H40 diagnosis at the Wilmer Eye Institute between 2013 and 2025 with at least five years of follow-up from the initial VF and at least five reliable 24-2 VFs within the first five years. All VFs were measured using Swedish Interactive Thresholding Algorithm (SITA)-Standard, SITA-Fast or SITA-Faster test strategies. Reliable VFs were defined as having <15% false positives and <25% false negatives where available for eyes with suspect, mild, or moderate glaucoma status or <50% false negatives for eyes with severe glaucoma^14^.

Each eye was also required to have at least one reliable Zeiss Cirrus Optical Coherence Tomography (OCT) scan within one year of the initial VF to evaluate the impact of OCT-derived metrics on rapid progression. Reliable OCTs were defined as having a minimum signal strength of six and superior, inferior, and average retinal nerve fiber layer (RNFL) thicknesses greater than 30um. RNFL thickness less than 30um is likely due to errors in registration or artifact, as it is approximately three standard deviations below the floor RNFL value on a Zeiss Cirrus OCT scan^15^.

Eyes were classified as having mild, moderate, or severe glaucoma based on their initial VF using Hodapp-Parish Anderson criteria for MD: mild (MD > -6dB), moderate (MD ≤ -6 dB and MD > -12 dB) and severe (MD ≤ -12 dB). A linear regression model was fit to the MD values for each eye to estimate the rate of MD progression. Patients were classified as rapid progressors if the rate of MD progression within the first five years was less than -1 dB/year and classified as non-rapid progressors otherwise.

Visual acuities for patients were measured using a Snellen chart at the first visit and converted to the logarithm of the minimum angle of resolution (logMAR). In line with previous literature, we converted “Count Fingers” to logMAR 1.85, “Hand Motion” to logMAR 2.30, “Light Perception” to logMAR 2.70 and “No Light Perception” to logMAR 3.00^16^.

Patients were classified as having received either aggressive procedures or minimally invasive procedures if they had a corresponding Current Procedural Terminology (CPT) code recorded within seven years of their first visual field assessment and glaucoma diagnosis. This would be during or after the period of rapid progression as previously defined for all included patients. Treatment codes for aggressive procedures included filtering surgeries or external ciliodestruction, whereas minimally invasive procedure codes included codes which corresponded to Minimally Invasive Glaucoma Surgery (MIGS) or laser procedures. Patients were assumed to be treated medically otherwise. A comprehensive list of CPT codes for aggressive and minimally invasive procedures is provided in online-only supplemental material.

### Socioeconomic Vulnerability Index

We used the Centers for Disease Control and Prevention and Agency for Toxic Substances and Disease Registry Social Vulnerability Index 2022 to estimate social vulnerability. This dataset provides an estimation of the overall socioeconomic vulnerability index (SVI) at a census tract level based on 1) socioeconomic status, 2) household characteristics, 3) racial and minority status, and 4) housing type and transportation. SVI ranges from 0 to 1 with a higher value representing a higher level (worse) of socioeconomic vulnerability. Patient ZIP codes were mapped to census tracts using the U.S. Department of Housing and Urban Development’s ZIP Code Crosswalk Files to assign an SVI to each patient

### Descriptive Statistical Analysis

First, we assessed whether there were any statistically significant differences between rapid progressors and non-rapid progressors for age, gender, SVI, race, and a variety of baseline measures from the first clinical visit, including MD, average RNFL thickness, cup-to-disc ratio, rim area, intraocular pressure, and visual acuity. The Wilcoxon rank sum test was used for continuous variables — all the continuous variables violated assumptions of normality (p < 0.05) based on a Shapiro-Wilk test — while the Chi-squared test was used for categorical variables.

We then structured our analysis to compare treatment patterns across two key dimensions: (1) progression status (rapid vs. non-rapid progressors), and (2) treatment intensities (aggressive, minimally invasive or medical) to create six distinct clinical groups for comparison (all possible combinations of progression status and treatment intensity). We used the Kruskal-Wallis test for continuous variables and the Chi-squared test used for categorical variables to assess differences across all six clinical groups. If any significant differences were found, post-hoc pairwise comparisons with Bonferroni correction were performed to determine which groups maintained significant differences.

### Modelling the impact of clinical, demographic and SVI variables on procedure selection

We utilized a multivariable logistic regression model to assess the extent to which clinical and demographic variables predict the likelihood of receiving aggressive procedures, minimally invasive procedures, or “any procedure” (i.e., aggressive or minimally invasive procedures). Separate analyses were conducted for rapid and non-rapid progressors to identify factors significantly associated with procedure selection decisions in each group. A sensitivity analysis was performed across all models by varying the intervention timeframe, adjusting the time frame for treatment receipt from seven years to alternative thresholds of three and five years. All statistical analysis was conducted using Python 3.9.0 with the Statsmodels 0.14.4 and Scipy 1.14.1 libraries.

## Results

A total of 2,782 eyes from 1,812 patients were included in this study. Table 1 lists key demographic and clinical characteristics of rapid (n = 192 eyes, 6.9%) and non-rapid progressors (n = 2,590 eyes, 93.1%) stratified by treatment received in the first seven years.

**Table 1.** Demographics.

|  | Overall | Rapid Progressors (n = 192, 6.9% of all eyes) |  |  |  | Non-Rapid Progressors (n = 2590, 93.1% of all eyes) |  |  |  | P-Value<br>(Comparison<br>between<br>overall rapid<br>and non-rapid<br>progressors) | P-Value<br>(Comparison<br>between<br>treatment<br>groups) |
| --- | --- | --- | --- | --- | --- | --- | --- | --- | --- | --- | --- |
|  |  | All<br>Treatments | Aggressive<br>Procedure | Minimally<br>Invasive<br>Procedure | Medical<br>Treatment | All<br>Treatments | Aggressive<br>Procedure | Minimally<br>Invasive<br>Procedure | Medical<br>Treatment |  |  |
| Patients (# eyes) | 2782 | 192 (100%) | 44 (22.9%) | 36 (18.8%) | 112 (58.3%) | 2590 (100%) | 152 (5.9%) | 286 (11.0%) | 2152 (83.1%) |  |  |
| Biological sex, n (%) |  |  |  |  |  |  |  |  |  | 0.69 | 0.58 |
| Female | 1509 (54.2%) | 101 (52.6%) | 20 (45.5%) | 16 (44.4%) | 65 (58.0%) | 1408 (54.4%) | 82 (53.9%) | 151 (52.8%) | 1175 (54.6%) |  |  |
| Male | 1273 (45.8%) | 91 (47.4%) | 24 (54.5%) | 20 (55.6%) | 47 (42.0%) | 1182 (45.6%) | 70 (46.1%) | 135 (47.2%) | 977 (45.4%) |  |  |
| Age, (years) | 64.00 [56.00, 71.00] | 67.00 [57.00, 73.25] | 71.00 [63.00, 74.00] | 67.00 [61.75, 74.25] | 65.00 [55.75, 72.25] | 64.00 [56.00, 71.00] | 66.00 [59.00, 72.25] | 66.50 [60.00, 71.00] | 64.00 [56.00, 71.00] | 0.01 | 0.00 |
| Race, n (%) |  |  |  |  |  |  |  |  |  | 0.30 | 0.06 |
| Asian | 213 (7.7%) | 14 (7.3%) | 4 (9.1%) | 3 (8.3%) | 7 (6.2%) | 199 (7.7%) | 9 (5.9%) | 18 (6.3%) | 172 (8.0%) |  |  |
| Black or African American | 906 (32.6%) | 78 (40.6%) | 16 (36.4%) | 8 (22.2%) | 54 (48.2%) | 828 (32.0%) | 55 (36.2%) | 77 (26.9%) | 696 (32.3%) |  |  |
| White or Caucasian | 1521 (54.7%) | 92 (47.9%) | 24 (54.5%) | 23 (63.9%) | 45 (40.2%) | 1429 (55.2%) | 77 (50.7%) | 174 (60.8%) | 1178 (54.7%) |  |  |
| Other | 101 (3.6%) | 6 (3.1%) | 0 (0.0%) | 2 (5.6%) | 4 (3.6%) | 95 (3.7%) | 8 (5.3%) | 9 (3.1%) | 78 (3.6%) |  |  |
| American Indian/Alaska Native | 6 (0.2%) | 0 (0.0%) | 0 (0.0%) | 0 (0.0%) | 0 (0.0%) | 6 (0.2%) | 0 (0.0%) | 2 (0.7%) | 4 (0.2%) |  |  |
| Other Pacific Islander | 8 (0.3%) | 1 (0.5%) | 0 (0.0%) | 0 (0.0%) | 1 (0.9%) | 7 (0.3%) | 2 (1.3%) | 0 (0.0%) | 5 (0.2%) |  |  |
| Not Disclosed/Unknown | 27 (1.0%) | 1 (0.5%) | 0 (0.0%) | 0 (0.0%) | 1 (0.9%) | 26 (1.0%) | 1 (0.7%) | 6 (2.1%) | 19 (0.9%) |  |  |
| SVI | 0.34 [0.22, 0.50] | 0.36 [0.23, 0.50] | 0.37 [0.22, 0.57] | 0.30 [0.21, 0.42] | 0.36 [0.25, 0.51] | 0.34 [0.22, 0.50] | 0.42 [0.23, 0.54] | 0.32 [0.20, 0.47] | 0.33 [0.22, 0.50] | 0.32 | 0.10 |
| VF Characteristics |  |  |  |  |  |  |  |  |  |  |  |
| MD at Baseline | -2.60 [-5.48, -0.79] | -4.36 [-8.63, -2.04] | -7.93 [-12.02, -4.21] | -3.59 [-7.58, -2.31] | -3.83 [-6.68, -1.60] | -2.52 [-5.31, -0.73] | -6.02 [-12.06, -2.67] | -2.70 [-5.39, -0.83] | -2.33 [-4.92, -0.64] | 0 | 0 |
| Slope of MD | -0.04 [-0.36, 0.26] | -1.41 [-1.97, -1.16] | -1.48 [-1.90, -1.17] | -1.34 [-1.68, -1.16] | -1.43 [-1.99, -1.17] | 0.00 [-0.28, 0.28] | -0.20 [-0.50, 0.19] | -0.03 [-0.40, 0.24] | 0.02 [-0.25, 0.30] | 0 | 0 |
| Number of Visits in five year duration | 6.00 [5.00, 7.00] | 6.00 [5.00, 8.00] | 6.00 [5.75, 8.00] | 6.00 [5.00, 8.25] | 6.00 [5.00, 8.00] | 6.00 [5.00, 7.00] | 6.00 [5.00, 8.00] | 6.00 [5.00, 7.00] | 6.00 [5.00, 7.00] | 0 | 0 |
| Clinical Characteristics |  |  |  |  |  |  |  |  |  |  |  |
| IOP (mmHg) | 17.00 [14.00–20.00] | 17.00 [14.00–21.00] | 17.00 [13.75–22.00] | 17.00 [14.00–22.00] | 16.00 [14.00–20.00] | 17.00 [14.00–20.00] | 18.00 [15.00–22.00] | 18.00 [15.00–22.00] | 17.00 [14.00–20.00] | 0.89 | 0 |
| Visual Acuity (logMAR) | 0.10 [0.00–0.18] | 0.14 [0.10–0.30] | 0.18 [0.10–0.42] | 0.10 [0.00–0.30] | 0.18 [0.10–0.30] | 0.10 [0.00–0.18] | 0.10 [0.00–0.30] | 0.10 [0.00–0.18] | 0.10 [0.00–0.18] | 0 | 0 |
| Glaucoma Severity |  |  |  |  |  |  |  |  |  | 0 | 0 |
| Mild Glaucoma | 2170 (78.0%) | 121 (63.0%) | 18 (40.9%) | 24 (66.7%) | 79 (70.5%) | 2049 (79.1%) | 76 (50.0%) | 231 (80.8%) | 1742 (80.9%) |  |  |
| Moderate Glaucoma | 396 (14.2%) | 46 (24.0%) | 15 (34.1%) | 8 (22.2%) | 23 (20.5%) | 350 (13.5%) | 36 (23.7%) | 38 (13.3%) | 276 (12.8%) |  |  |
| Severe Glaucoma | 216 (7.8%) | 25 (13.0%) | 11 (25.0%) | 4 (11.1%) | 10 (8.9%) | 191 (7.4%) | 40 (26.3%) | 17 (5.9%) | 134 (6.2%) |  |  |
| OCT Characteristics |  |  |  |  |  |  |  |  |  |  |  |
| Average RNFL Thickness (µm) | 78.55 [69.02, 87.05] | 74.56 [63.48, 84.25] | 68.93 [56.93, 81.10] | 65.94 [59.91, 77.04] | 78.38 [69.75, 85.59] | 78.78 [69.46, 87.40] | 68.72 [60.84, 79.30] | 75.00 [66.96, 82.41] | 79.68 [70.83, 88.37] | 0 | 0 |
| Average Cup Disc Ratio | 0.67 [0.57, 0.74] | 0.71 [0.59, 0.79] | 0.76 [0.63, 0.79] | 0.73 [0.66, 0.80] | 0.69 [0.57, 0.77] | 0.66 [0.56, 0.74] | 0.73 [0.62, 0.81] | 0.67 [0.57, 0.73] | 0.66 [0.56, 0.73] | 0 | 0 |
| RIM Area (mm²) | 1.01 [0.83, 1.21] | 0.94 [0.74, 1.19] | 0.81 [0.60, 1.03] | 0.86 [0.68, 0.98] | 1.05 [0.81, 1.30] | 1.01 [0.84, 1.21] | 0.80 [0.67, 1.10] | 0.97 [0.81, 1.14] | 1.03 [0.85, 1.23] | 0.05 | 0 |

\*Continuous variables are presented as median [IQR].

We first examined differences in demographic and social vulnerability characteristics between rapid and non-rapid glaucoma progressors, emphasizing factors potentially influencing treatment decisions (Table 1). Rapid progressors were older than non-rapid progressors (median 67 [57–73] vs 64 [56–71] years, p = 0.01). No significant differences emerged in biological sex (p = 0.69), racial distribution (p = 0.30), or Social Vulnerability Index (SVI, p = 0.32) (see Table 1 for full details).

Regarding baseline clinical characteristics, rapid progressors demonstrated significantly worse glaucoma severity distribution (p < 0.001), characterized by greater proportions of moderate (24.0% vs. 13.5%) and severe glaucoma cases (13.0% vs. 7.4%). Rapid progressors and non-rapid progressors had the same median number of visits (6.0), though the distribution was significantly different (p < 0.001). Subsequent analysis of the distributions shows that the non-rapid progressor group had a more pronounced tail than the rapid-progressor group (higher kurtosis), reflecting more outliers in visit frequencies. Additionally, rapid progressors had significantly worse baseline visual field mean deviation (MD: -4.36 vs. -2.52 dB, p < 0.001), poorer visual acuity (logMAR: 0.14 vs. 0.10, p < 0.001), and worse optical coherence tomography (OCT) measures, including thinner retinal nerve fiber layer (RNFL) thickness (74.6 µm vs. 78.8 µm, p < 0.001) and larger cup-to-disc ratios (0.71 vs. 0.66, p < 0.001). No significant differences were identified in intraocular pressure at first clinic visit (IOP) (see Table 1 for full details).

When we examined differences between the six progression-treatments groups (rapid/non-rapid progressors and by treatment type), we found significant differences in all measured variables except biological sex (p=0.58), race (p = 0.06), and SVI (p=0.10). Subsequent pairwise analyses identified significant differences in MD slope for each treatment type between rapid vs. non-rapid progressors (all p<0.001). Treatment patterns followed disease severity at initial presentation. Rapid progressors receiving aggressive procedures had significantly worse baseline MD (−7.93 [−12.02, −4.21] dB) compared to all other groups (range of medians: −2.33 to −6.02 dB, all p < 0.001). Among non-rapid progressors, those with a worse baseline MD were more likely to receive aggressive procedures (p < 0.001). Rapid progressors managed medically required significantly more follow-up visits than non-rapid medical patients (p < 0.001), indicating closer monitoring despite conservative management. Similarly, among non-rapid progressors, patients receiving aggressive procedures attended more visits compared to minimally invasive or medical groups (both p ≤ 0.020). Rapid progressors who received aggressive procedures had significantly worse visual acuity (logMAR 0.31 vs. 0.14; p=0.004) and thinner RNFL thickness (medians 68.93 µm vs. 79.68 µm; p≤0.003) compared to non-rapid progressors who received minimally invasive procedures or medical treatment, highlighting worse baseline visual impairment and structural damage. Likewise, among non-rapid progressors, aggressive procedures remained associated with poorer visual acuity and thinner RNFL compared with minimally invasive or medical treatments (all p < 0.001), indicating that initial severity is associated with aggressive procedures regardless of rapid progression status.

Next, we assessed which demographic, clinical, and socioeconomic factors were associated with different intervention types (aggressive, minimally invasive, or any procedure) using logistic regression, first across all patients (Table 2), then rapid progressors (Table 3), and finally non-rapid progressors (Table 4).

**Table 2.** Logistic regression for predictors of intervention (aggressive procedures, minimally invasive procedures, or any procedure) in all patients.

| Variable | Aggressive Procedures Odds Ratio (95% CI) | P-value | Minimally Invasive Procedures Odds Ratio (95% CI) | P-value | Any Procedure Odds Ratio (95% CI) | P-value |
| --- | --- | --- | --- | --- | --- | --- |
| Age (per 1 year increase) | 1.01 (1.00–1.02) | 0.206 | 1.02 (1.01–1.03) | 0.000* | 1.02 (1.01–1.03) | 0.000* |
| Gender (Male vs. Female) | 0.96 (0.70–1.31) | 0.805 | 0.99 (0.78–1.25) | 0.909 | 0.97 (0.80–1.19) | 0.799 |
| Race (Asian vs. White) | 0.91 (0.48–1.72) | 0.777 | 0.93 (0.57–1.52) | 0.766 | 0.88 (0.58–1.33) | 0.534 |
| Race (Black or African American vs. White) | 1.07 (0.72–1.59) | 0.730 | 0.85 (0.62–1.16) | 0.298 | 0.91 (0.70–1.18) | 0.455 |
| Race (Other vs. White) | 1.13 (0.51–2.49) | 0.757 | 1.00 (0.52–1.94) | 0.994 | 1.02 (0.60–1.76) | 0.936 |
| SVI (per unit increase) | 1.63 (0.66–4.04) | 0.292 | 0.65 (0.31–1.35) | 0.246 | 0.91 (0.50–1.67) | 0.768 |
| Rapid Progressor Status | 3.83 (2.56–5.74) | 0.000* | 1.82 (1.22–2.71) | 0.003* | 3.15 (2.28–4.35) | 0.000* |
| MD at First Visit (per dB decrease) | 1.10 (1.06–1.12) | 0.000* | 0.95 (0.93–0.99) | 0.004* | 1.03 (1.00–1.05) | 0.024* |
| IOP (per mmHg increase) | 1.04 (1.01–1.07) | 0.008* | 1.06 (1.04–1.08) | 0.000* | 1.06 (1.04–1.08) | 0.000* |
| VA (per unit logMAR increase) | 1.73 (1.03–2.92) | 0.04* | 0.63 (0.34–1.15) | 0.134 | 1.08 (0.70–1.67) | 0.719 |
| Average Cup Disc Ratio (per unit increase) | 0.60 (0.13–2.75) | 0.507 | 0.66 (0.23–1.86) | 0.427 | 0.60 (0.24–1.48) | 0.265 |
| Average RNFL Thickness (per $\mu\text{m}$ increase) | 0.99 (0.98–1.01) | 0.227 | 0.98 (0.97–0.99) | 0.000* | 0.98 (0.97–0.99) | 0.000* |
| Rim Area (per $\text{mm}^2$ increase) | 0.25 (0.10–0.60) | 0.002* | 0.55 (0.29–1.04) | 0.067 | 0.36 (0.21–0.63) | 0.000* |

\* Females serve as the reference group for gender, and white patients serve as the reference group for race. All procedure groups are compared with those not in the procedure as a reference group. Asterisks indicate statistically significant p-values ( $p < 0.05$ )

**Table 3.** Logistic regression for predictors of intervention (aggressive procedures, minimally invasive procedures, or any procedure) in rapid progressors.

| Variable | Aggressive Procedures Odds Ratio (95% CI) | P-value | Minimally Invasive Procedures Odds Ratio (95% CI) | P-value | Any Procedure Odds Ratio (95% CI) | P-value |
| --- | --- | --- | --- | --- | --- | --- |
| Age (per 1 year increase) | 1.02 (0.99–1.06) | 0.201 | 1.00 (0.96–1.03) | 0.882 | 1.01 (0.98–1.04) | 0.515 |
| Gender (Male vs. Female) | 1.51 (0.72–3.20) | 0.276 | 1.70 (0.75–3.87) | 0.204 | 1.94 (0.99–3.80) | 0.054 |
| Race (Asian vs. White) | 1.51 (0.39–5.88) | 0.553 | 0.82 (0.19–3.53) | 0.786 | 1.08 (0.31–3.76) | 0.91 |
| Race (Black or African American vs. White) | 0.79 (0.30–2.06) | 0.633 | 0.49 (0.17–1.38) | 0.176 | 0.55 (0.24–1.25) | 0.151 |
| Race (Other vs. White) | 0.00 (0.00–inf) | 0.995 | 1.35 (0.21–8.75) | 0.756 | 0.50 (0.08–3.20) | 0.461 |
| SVI (per unit increase) | 1.99 (0.19–20.29) | 0.563 | 0.05 (0.00–0.76) | 0.031* | 0.25 (0.03–1.92) | 0.182 |
| MD at First Visit (per dB decrease) | 1.10 (1.02–1.18) | 0.015* | 0.93 (0.85–1.03) | 0.159 | 1.04 (0.97–1.11) | 0.215 |
| IOP (per mmHg increase) | 1.03 (0.96–1.11) | 0.383 | 1.08 (0.99–1.17) | 0.077 | 1.07 (1.00–1.14) | 0.036* |
| VA (per unit logMAR increase) | 1.61 (0.54–4.81) | 0.39 | 0.35 (0.05–2.24) | 0.265 | 0.97 (0.33–2.82) | 0.952 |
| Average Cup Disc Ratio (per unit increase) | 0.37 (0.01–19.06) | 0.618 | 6.85 (0.06–754.67) | 0.422 | 1.28 (0.04–39.98) | 0.888 |
| Average RNFL Thickness (per $\mu\text{m}$ increase) | 1.02 (0.98–1.05) | 0.334 | 0.99 (0.95–1.02) | 0.506 | 1.01 (0.98–1.03) | 0.71 |
| Rim Area (per $\text{mm}^2$ increase) | 0.14 (0.02–1.16) | 0.069 | 0.29 (0.03–2.77) | 0.285 | 0.10 (0.02–0.62) | 0.013* |

\* Females serve as the reference group for gender, and white patients serve as the reference group for race. All procedure groups are compared with those not in the procedure as a reference group. Asterisks indicate statistically significant p-values ( $p < 0.05$ )

**Table 4.** Logistic regression for predictors of intervention (aggressive procedures, minimally invasive procedures, or any procedure) in non-rapid progressors.

| Variable | Aggressive Procedures Odds Ratio (95% CI) | Aggressive Procedures P-value | Minimally Invasive Procedures Odds Ratio (95% CI) | Minimally Invasive Procedures P-value | Any Procedure Odds Ratio (95% CI) | Any Procedure P-value |
| --- | --- | --- | --- | --- | --- | --- |
| Age (per 1 year increase) | 1.01 (0.99–1.02) | 0.407 | 1.02 (1.01–1.04) | 0.000* | 1.02 (1.01–1.03) | 0.000* |
| Gender (Male vs. Female) | 0.89 (0.63–1.25) | 0.49 | 0.95 (0.74–1.23) | 0.706 | 0.92 (0.74–1.14) | 0.453 |
| Race (Asian vs. White) | 0.78 (0.37–1.64) | 0.519 | 0.93 (0.55–1.57) | 0.773 | 0.84 (0.53–1.31) | 0.431 |
| Race (Black or African American vs. White) | 1.14 (0.74–1.77) | 0.543 | 0.86 (0.62–1.20) | 0.391 | 0.95 (0.72–1.25) | 0.692 |
| Race (Other vs. White) | 1.39 (0.62–3.10) | 0.419 | 0.94 (0.46–1.94) | 0.874 | 1.10 (0.63–1.94) | 0.733 |
| SVI (per unit increase) | 1.50 (0.56–4.07) | 0.422 | 0.78 (0.36–1.67) | 0.518 | 0.97 (0.51–1.83) | 0.913 |
| MD at First Visit (per dB decrease) | 1.10 (1.06–1.12) | 0.000* | 0.96 (0.93–0.99) | 0.011* | 1.02 (1.00–1.05) | 0.045* |
| IOP (per mmHg increase) | 1.04 (1.01–1.07) | 0.008* | 1.06 (1.04–1.09) | 0.000* | 1.06 (1.04–1.08) | 0.000* |
| VA (per unit logMAR increase) | 1.73 (0.94–3.20) | 0.079 | 0.73 (0.38–1.43) | 0.365 | 1.13 (0.70–1.82) | 0.616 |
| Average Cup Disc Ratio (per unit increase) | 0.69 (0.13–3.70) | 0.662 | 0.60 (0.21–1.75) | 0.351 | 0.60 (0.23–1.54) | 0.289 |
| Average RNFL Thickness (per μm increase) | 0.98 (0.97–1.00) | 0.073 | 0.98 (0.96–0.99) | 0.000* | 0.98 (0.97–0.99) | 0.000* |
| Rim Area (per mm<sup>2</sup> increase) | 0.30 (0.11–0.79) | 0.015* | 0.61 (0.31–1.20) | 0.151 | 0.44 (0.25–0.78) | 0.005* |

\* Females serve as the reference group for gender, and white patients serve as the reference group for race. All procedure groups are compared with those not in the procedure as a reference group. Asterisks indicate statistically significant p-values ( $p < 0.05$ ).

In the overall patient cohort (Table 2), demographic and socioeconomic factors played only a minor role in surgical decision-making. Gender, race, and Social Vulnerability Index were not statistically associated with any category of intervention (all p > 0.05). Age exerted a small yet significant effect as each additional year increased the odds of undergoing a minimally invasive procedure by 2% (odds ratio [OR]: 1.02 per year, 95% confidence interval [CI] 1.01–1.03, p < 0.001) and the odds of receiving any procedure by the same margin (OR: 1.02 per year, 95% CI: 1.01–1.03, p < 0.001), but had no impact on aggressive surgery (p = 0.206).

Among clinical predictors, rapid progression strongly increased the odds of receiving aggressive procedures (OR 3.83, 95 % CI 2.56–5.74, p < 0.001), minimally invasive surgery (OR 1.82, 95 % CI 1.22–2.71, p = 0.003) and any procedure (OR 3.15, 95 % CI 2.28–4.35, p < 0.001). Worse visual field damage at baseline, as indicated by more negative MD, also predicted aggressive procedures (OR 1.10, 95 % CI 1.06–1.12, p < 0.001) and any procedure (OR 1.03, 95 % CI 1.00–1.05, p = 0.024) but decreased the odds of minimally invasive surgery (OR 0.95, 95 % CI 0.93–0.99, p = 0.004), suggesting that eyes with milder functional damage are preferentially offered lower-risk MIGS. Elevated intraocular pressure (IOP) at the first clinical visit was a consistent predictor across all intervention types—aggressive (OR: 1.04 per mmHg increase, 95 % CI: 1.01–1.07, p = 0.008), minimally invasive (OR: 1.06 per mmHg increase, 95 % CI: 1.04–1.08, p < 0.001), and any procedure (OR: 1.06 per mmHg increase, 95 % CI: 1.04–1.08, p < 0.001). Worse visual acuity (higher logMAR) significantly increased the likelihood of aggressive interventions (OR: 1.73 per unit increase, 95 % CI: 1.03–2.92, p = 0.040). On structural evaluation, thinner average retinal nerve fiber layer (RNFL) thickness was significantly associated with greater use of minimally invasive procedures and with any procedure (OR 0.98 per µm, 95 % CI 0.97– 0.99, p < 0.001 for both). Larger rim area were significantly associated with lower odds of aggressive procedures (OR: 0.25 per mm² increase, 95% CI: 0.10–0.60, p = 0.002) and any procedure (OR 0.36 per mm² increase, 95% CI 0.21–0.63, p < 0.001). Average cup-to-disc ratio did not significantly influence procedural likelihood.

Among rapid progressors (Table 3), we found that key predictors from the overall patient cohort largely remained consistent except for SVI. Elevated IOP at the first clinical visit continued to significantly increase the likelihood of receiving any procedure (OR 1.07 per mmHg increase, 95% CI: [1.00, 1.14], p < 0.05). Worse initial MD significantly correlated with higher odds of aggressive procedures (OR 1.10 per dB decrease, 95% CI: 1.02–1.18, p = 0.015). Larger rim area significantly reduced the odds of any procedure (OR 0.10 per mm² increase, 95% CI 0.02–0.62, p = 0.013) and demonstrated a similar, though non-significant, trend for aggressive surgery (OR 0.14 per mm² increase, 95% CI 0.02-1.16, p = 0.069).

A novel and clinically important finding was that higher SVI significantly reduced the likelihood of minimally invasive procedures (OR 0.05 per unit increase, 95% CI: 0.00–0.76, p = 0.031), suggesting socioeconomic factors might limit access to minimally invasive options among rapid progressors. Higher SVI also tended to be associated with aggressive procedures in this cohort, although this did not reach statistical significance (OR 1.99 per unit increase, 95% CI: 0.19–20.29, p = 0.563). Demographic factors such as age, sex, and race, as well as baseline visual acuity and average cup-to-disc ratio did not reach statistical significance.

Among non-rapid progressors (Table 4), predictors of intervention were also similar to the overall cohort but varied in magnitude. Age was significantly associated with higher odds of minimally invasive procedures (OR 1.02 per year, 95% CI 1.01–1.04, p < 0.001) and any procedure (OR 1.02 per year, 95% CI 1.01–1.03, p < 0.001). Worse baseline MD (more negative) was significantly associated with greater odds of aggressive procedures (OR 1.10 per dB decrease, 95 % CI 1.06–1.12, p < 0.001) and of any procedure (OR 1.02, 95 % CI 1.00–1.05, p = 0.045), while modestly lowering the odds of minimally invasive surgery (OR 0.96, 95 % CI 0.93–0.99, p = 0.011). Higher initial IOP significantly predicted intervention, increasing the odds of aggressive (OR 1.04 per mmHg, 95% CI 1.01–1.07, p = 0.008), minimally invasive (OR 1.06, 95% CI 1.04–1.09, p < 0.001), and any procedure (OR 1.06, 95% CI 1.04– 1.08, p < 0.001). Thinner retinal nerve fiber layer thickness was significantly associated with higher odds of minimally invasive (OR 0.98 per µm, 95% CI 0.96–0.99, p < 0.001) and any procedure (OR 0.98, 95% CI 0.97–0.99, p < 0.001). No significant differences were observed for sex, race, SVI, baseline visual acuity, and average cup-to-disc ratio.

### Sensitivity Analysis

When shortening the timeframe for treatment after initial clinical diagnosis from seven years to five and three years, some predictors changed in their statistical significance. Age was a significant predictor in the seven-year analysis for minimally invasive procedures and any procedure (e.g., seven-year odds ratio = 1.02, p < 0.001). However, they often lost statistical significance in the three-year models (e.g., any procedure three-year odds ratio = 1.02, p = 0.06).

In contrast, elevated baseline IOP showed a consistent positive association with the likelihood of aggressive, minimally invasive, and any procedures. For example, each additional mmHg of IOP increased the odds of aggressive procedures by roughly 4–5% in both 3-year (OR = 1.05, p = 0.006) and 5-year models (OR = 1.05, p < 0.001), and maintained similar associations at 7 years (OR = 1.04, p = 0.008). Therefore, despite narrowing the treatment window, baseline IOP and key functional and structural measures of disease severity (worse MD, smaller rim area) remained the most robust predictors of both early and later interventions.

Finally, the primary predictors of aggressive procedures, particularly disease severity, remained consistent across all models and stayed statistically significant across all timeframes. For instance, a better mean deviation at the first visit continued to be a strong predictor of lower odds of aggressive procedures (e.g., OR = 1.09, 95% CI: 1.05–1.12 at 3 years; OR = 1.09, 95% CI: 1.05–1.12 at 5 years; OR = 1.10, 95% CI: 1.06–1.12 at 7 years, all p < 0.001). Similarly, the optic disc rim area consistently demonstrated large effect sizes for predicting aggressive procedures (e.g., OR ≈ 0.16–0.25 across models, p < 0.004), indicating that patients with smaller rim areas were more likely to undergo early and more invasive procedures.

## Discussion

In this study, we evaluated the clinical and socioeconomic determinants of treatment decisions in glaucoma patients with different visual field progression rates. Despite finding that rapid progression was associated with greater odds of receiving either aggressive procedures (OR 3.83) or any procedure (OR 3.15), only 42% of rapidly progressive patients underwent any non-medical procedure, of which 23% were aggressive IOP lowering procedures within seven years of their initial visit. Among patients who progressed rapidly, higher baseline intraocular pressures increased the odds of any procedure, while worse functional or structural parameters increased the overall likelihood of aggressive procedures.

Furthermore, a higher social vulnerability index was associated with a lower likelihood of receiving less aggressive therapy, which highlights the potential role of socioeconomic factors in shaping treatment decisions.

While previous studies^3,4^ have reported a 5-12% rapid progression in glaucomatous patients, to our knowledge, this is the first study that specifically reports treatment patterns in rapid progressors and how these may vary from non-rapid progressors. In our cohort of 2,782 eyes from 1,812 patients, only 23% of rapidly progressing eyes ultimately underwent aggressive procedures within seven years of diagnosis. We also found that there was no significant pairwise difference for MD slope between treatment groups for rapid progressors – in other words, if there was a faster rate of MD worsening was not associated with more aggressive treatment within the rapid progressor cohort. This relatively low percentage and similar MD slopes between treatment categories for rapid progressors is clinically significant and may not be intuitive, as rapid progression typically raises clinical concerns warranting aggressive interventions to preserve visual function. Potential reasons for this observed gap include clinician reliance on baseline disease severity markers (e.g., intraocular pressure, structural OCT findings) over functional progression rates, patient hesitancy, or refusal of invasive procedures, perceived surgical risks, socioeconomic barriers limiting access to interventions, or clinician preference for conservative management due to concerns about complications. Specifically, in the overall cohort and rapid progressors, eyes with a higher IOP, worse MD, smaller optic disc rim area or thinner RNFL at presentation were significantly more likely to undergo aggressive procedures or any procedure. When we restricted the analysis to rapid progressors alone, the association between structural and functional damage as a predictor of aggressive procedures remained robust. Our data suggest that rapid progression, as defined retrospectively from at least five VF tests, may not always align with initial clinical impressions or decisions which may be based on baseline disease severity or IOP. Since clinicians may not have access to longitudinal VF progression data when initially deciding on treatment escalation, our results underscore the potential value of incorporating standardized and timely measures of VF loss rate earlier in clinical decision-making processes to better identify and manage patients who may rapidly progress or incorporating models which can forecast rapid progression risk from early data points^17, 18^. This observation highlights a potential gap in aggressive intervention among patients demonstrating rapid functional decline, as most rapidly progressing patients did not receive definitive aggressive procedures. Future studies could prospectively assess how earlier identification and consideration of rapid VF progression rates could influence treatment escalation decisions in clinical practice.

Across the rapid progressor population, patients with worse (higher) SVI who were rapid progressors had a markedly reduced likelihood of receiving minimally invasive procedures (OR = 0.05, p = 0.031). This may reflect more advanced disease at presentation or reduced access to healthcare in earlier stages. Prior research has consistently found that higher neighborhood-level socioeconomic strongly correlates with delayed diagnosis or inconsistent follow-up, which may lead to more urgent need for definitive procedures once severe damage is detected^8, 9, 10, 12^. Although we adjusted our analysis for baseline disease severity, our results also highlight a significant association between socioeconomic vulnerability and the types of glaucoma interventions patients received, underscoring potential disparities in access or utilization of different treatment modalities. This suggests potential unmeasured confounding variables, from disease severity factors associated with SVI as well as a possible direct causal impact of SVI on treatment selection. These findings align with previous literature demonstrating disparities in access to minimally invasive procedures among different demographic groups^19^.

In contrast with previous literature, we find that demographic characteristics beyond age do not shape treatment approaches^8,12,20^. We find that an increase in age was associated with a higher odds of minimally invasive procedures (OR = 1.02 per year, p < 0.001) or any procedure (OR = 1.02, p < 0.001). However, neither gender nor race showed an independent relationship with treatment selection. This divergence from earlier work may reflect our adjustment for socioeconomic vulnerability using SVI, a determinant that previous analyses did not include.

There are several limitations to this study. This was a retrospective, single-center study in an academic setting, which may limit the generalizability of the results to other practice environments or diverse patient populations. Treatment decisions and outcomes were drawn from clinical data analyzed retrospectively, and unmeasured confounders such as patient preferences, medication adherence, and clinician and clinic-specific practice patterns may have influenced both the selection of therapy and observed progression rates, especially when considering the effect of socioeconomic status. We also relied on administrative CPT and ICD-10 codes to capture both procedures and diagnoses, which may have possible misclassifications, and repeated interventions in the seven-year follow-up period post initial diagnoses were not explicitly accounted for. Furthermore, the SVI used in this study provides a census-tract level estimate of socioeconomic status which is necessarily a coarse measure, and may therefore not have fully encapsulated the individual level social determinants of health that can affect treatment access and decision-making. Finally, our inclusion criteria of requiring at least five reliable visual fields within five years and one reliable OCT within one year of first visual field may have excluded patients with more severe disease, limited follow-up or suboptimal testing, which may have introduced selection bias into our results, as such patients may have rapidly progressed had they not had a procedure earlier in their disease course.

## Conclusion

In conclusion, this study demonstrates that although rapid visual field progression was associated with greater likelihood of receiving aggressive or minimally invasive treatments, fewer than one in four rapidly progressing eyes received aggressive intervention within seven years of initial presentation. Furthermore, our data suggest that socioeconomic factors play a pivotal role, especially in the reduced uptake of minimally invasive treatment in rapidly progressing patients with a worse SVI, highlighting the need to address disparities in healthcare access and utilization. Future research efforts should focus on refining risk stratification tools, potentially integrating the current or forecasted rate of VF progression more prominently into treatment algorithms and mitigating socioeconomic barriers so we can optimize outcomes for patients with rapidly progressing glaucoma.

## Data Availability

All data produced in the present study are available upon reasonable request to the authors

## Abbreviations and Acronyms

ADI: Area Deprivation Index
CI: Confidence Interval
CPT: Current Procedural Terminology
dB: Decibel
ICD: International Classification of Diseases
IOP: Intraocular Pressure
logMAR: Logarithm of the Minimum Angle of Resolution
MD: Mean Deviation
MIGS: Minimally Invasive Glaucoma Surgery
OCT: Optical Coherence Tomography
OR: Odds Ratio
RNFL: Retinal Nerve Fiber Layer
SITA: Swedish Interactive Thresholding Algorithm
SVI: Social Vulnerability Index
VA: Visual Acuity
VF: Visual Field
ZIP: Zone Improvement Plan (ZIP Code)

## References

1. Quigley HA, Broman AT. The number of people with glaucoma worldwide in 2010 and 2020. Br J Ophthalmol. 2006;90(3):262–267.

2. Tham Y-C, Li X, Wong TY, Quigley HA, Aung T, Cheng C-Y. Global prevalence of glaucoma and projections of glaucoma burden through 2040. Ophthalmology. 2014;121(11):2081–2090.

3. Jackson AB, Martin KR, Coote MA, et al. Fast progressors in glaucoma: prevalence based on global and central visual field loss. Ophthalmology. 2023;130(5):462–468.

4. Huang X, Poursoroush A, Sun J, et al. Identifying factors associated with fast visual field progression in patients with ocular hypertension based on unsupervised machine learning. J Glaucoma. 2024;33(11):815–822.

5. Chauhan BC, Garway-Heath DF, Goñi FJ, et al. Practical recommendations for measuring rates of visual field change in glaucoma. Br J Ophthalmol. 2008;92(4):569–573.

6. Kim JH, Rabiolo A, Morales E, et al. Risk factors for fast visual field progression in glaucoma. Am J Ophthalmol. 2019;207:268–278.

7. Prata TS, De Moraes CGV, Teng CC, et al. Factors affecting rates of visual field progression in glaucoma patients with optic disc hemorrhage. Ophthalmology. 2010;117(1):24–29.

8. Shaheen A, Medeiros FA, Swaminathan SS. Association between greater social vulnerability and delayed glaucoma surgery. Am J Ophthalmol. 2024;268:123–135.

9. Tseng VL, Kitayama K, Yu F, Coleman AL. Disparities in glaucoma surgery: a review of current evidence and future directions for improvement. Transl Vis Sci Technol. 2023;12(9):2.

10. Tseng VL, Kitayama K, Yu F, Pan D, Coleman AL. Social vulnerability, prevalence of glaucoma, and incidence of glaucoma surgery in the California Medicare population. Ophthalmol Glaucoma. 2023;6(6):616–625.

11. Almidani L, Bradley C, Herbert P, et al. The impact of social vulnerability on structural and functional glaucoma severity, worsening, and variability. Ophthalmol Glaucoma. 2024;7(4):380–390.

12. Musa I, Bansal S, Kaleem MA. Barriers to care in the treatment of glaucoma: socioeconomic elements that impact the diagnosis, treatment, and outcomes in glaucoma patients. Curr Ophthalmol Rep. 2022;10(3):85–90.

13. Folgar FA, De Moraes CGV, Prata TS, et al. Glaucoma surgery decreases the rates of localized and global visual field progression. Am J Ophthalmol. 2010;149(2):258–264.

14. Yohannan J, Cheng M, Da J, et al. Evidence-based criteria for determining peripapillary OCT reliability. Ophthalmology. 2020;127(2):167–176.

15. Yohannan J, Wang J, Brown J, et al. Evidence-based criteria for assessment of visual field reliability. Ophthalmology. 2017;124(11):1612–1620.

16. Schulze-Bonsel, K., Feltgen, N., Burau, H., Hansen, L., & Bach, M. (2006). Visual acuities “hand motion” and “counting fingers” can be quantified with the freiburg visual acuity test. Investigative ophthalmology & visual science, 47(3), 1236–1240.

17. Herbert P, Hou K, Bradley C, et al. Forecasting risk of future rapid glaucoma worsening using early visual field, OCT, and clinical data. Ophthalmol Glaucoma. 2023;6(5):466–473.

18. Shuldiner SR, Boland MV, Ramulu PY, et al. Predicting eyes at risk for rapid glaucoma progression based on an initial visual field test using machine learning. PLoS One. 2021;16(4):e0249856.

19. Vasu P, Hall RP, Wagner IV, et al. Racial disparities in microinvasive glaucoma surgery for management of primary open-angle glaucoma: a propensity-matched cohort study. Am J Ophthalmol. 2025;271:96–103.

20. Allison K, Hodges B, Shahid MM, Feng C. Racial and gender disparities for glaucoma treatment rates in Upstate New York. J Clin Med. 2024;13(23):7225.

